# Theoretical derivation and clinical validation of the resolution limit of human eye to spherical lens change:A Self-controlled Study

**DOI:** 10.1101/2022.04.13.22273857

**Authors:** Zhen Yi, Gao Jie, Cao Kai, Shen Jing, Zhang wei, Dai Yun

## Abstract

**Background/Aims:** To deduce theoretically and verify the resolution limit of human eye to spherical lens change for more reasonable design of the trial lenses.

**Methods:** First, the resolution limit of discernible change in spherical power was derived based on the optical model. Then, the volunteers were observed to see if they could perceive the changes in spherical power as per the resolution limit and compare the difference in the best corrected visual acuity obtained with the resolution limit and interval of 0.25D.

**Results:** Assuming that the cone cell diameter is 3 μm and the pupil diameter of 4 mm, the theoretically resolution limit was 0.05D. When the diopter of spherical power was increased, the ratios of ability to perceive 0.05D spherical lens change were 98.3% and 96.7% in right and left eyes. When the diopter of spherical power was decreased, the ratios of ability to perceive 0.05D spherical lens change were 78.9% and 83.2% in right and left eyes. The best corrected visual acuity obtained with the 0.05 D interval trial lens was significantly better than in the 0.25 D interval on both eyes (Right eye -0.04±0.07 vs -0.02±0.06, t=6.729, P<0.001; Left eye -0.07±0.06 vs -0.04±0.06, t=8.825, P<0.001).

**Conclusion:** The resolution limit of human eye to spherical lens change was about 0.05D and the better corrected visual acuity can be obtained by adjusting the spherical power at an interval of 0.05D.

**What is already known on this topic:** At present, the spherical power is generally adjusted at 0.25D for optometry. In clinical practice, we find that a more than 80% of myopia patients with clear red prototypes will directly see clear green in case of a decrease by 0.25D, unable to achieve the red-green balance.

**What this study adds:** More than 80% of myopia patients can perceive the changes in spherical power as per 0.05D.

**How this study might affect research, practice or policy:** Adjusting the spherical power per 0.05 D can help us to achieve a higher full correction rate (>80%) and realize better visual acuity.

**[Synopsis/Precis]:** The resolution limit of human eye to spherical lens change is about 0.05D and better corrected visual acuity can be obtained by adjusting the spherical power at an interval of 0.05D than 0.25D.

## Introduction

In the information age, the prevalence of myopia among adolescents is increasing year by year, evidenced by up to 80% of myopia among Chinese high school students. Frame glasses are still the main method for myopia correction. Previous studies have shown that overcorrection or undercorrection will lead to the accelerated progression of myopia. Spherical-power full correction is recommended for adolescent myopia^[1 2]^. The red-green Duochrome test is an important step in subjective refraction, and a method to determine the maximum plus to maximum visual acuity (MPMVA)^[3]^. At present, the spherical power is generally adjusted at 0.25D for optometry. In clinical practice, we find that a considerable proportion of patients with clear red prototypes will directly see clear green in case of a decrease by 0.25D, unable to achieve the red-green balance. In order to avoid overcorrection, the prescription of optometry often chooses the diopter of undercorrection with clear red prototypes.

From the middle of the 19th century to the early 20th century, the interval of spherical-power trial case lenses was reduced from 1D to 0.25D and has been used up to now. However, we have not found the theoretical basis for setting the interval at 0.25D, which may be related to the cost of lens manufacturing at that time. In order to determine the adjustment interval of spherical-power lens more scientifically and reasonably and help patients get better visual quality, this study firstly calculated the theoretically derived resolution limit of human eye to spherical lens change based on the optical model, and then observed the actual resolution values of volunteers to compare the two for any consistency.

## Materials and Methods

### Calculation of the theoretically derived resolution limit of human eye to spherical lens change

Let the diopter of a human eye optical model be A (D), the diameter of human cone cell B (μm), and the pupil diameter *x*(mm). As shown in Fig. 1, the parallel light passes through the optical model of human eye and forms a focus spot on the image plane (retina), exciting one cone cell only. When the spherical lens with diopter y is decreased in front of the model, the focus moves back to the retina, and the parallel light will form a diffuse spots in the retina. When the diameter of the diffuse spots is longer than or equal to B, two cone cells will be excited, and may thus be perceived. The following formulae can be derived from Fig. 1:

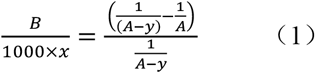

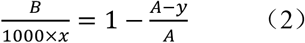

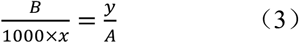

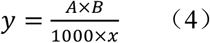

**Figure 1.**
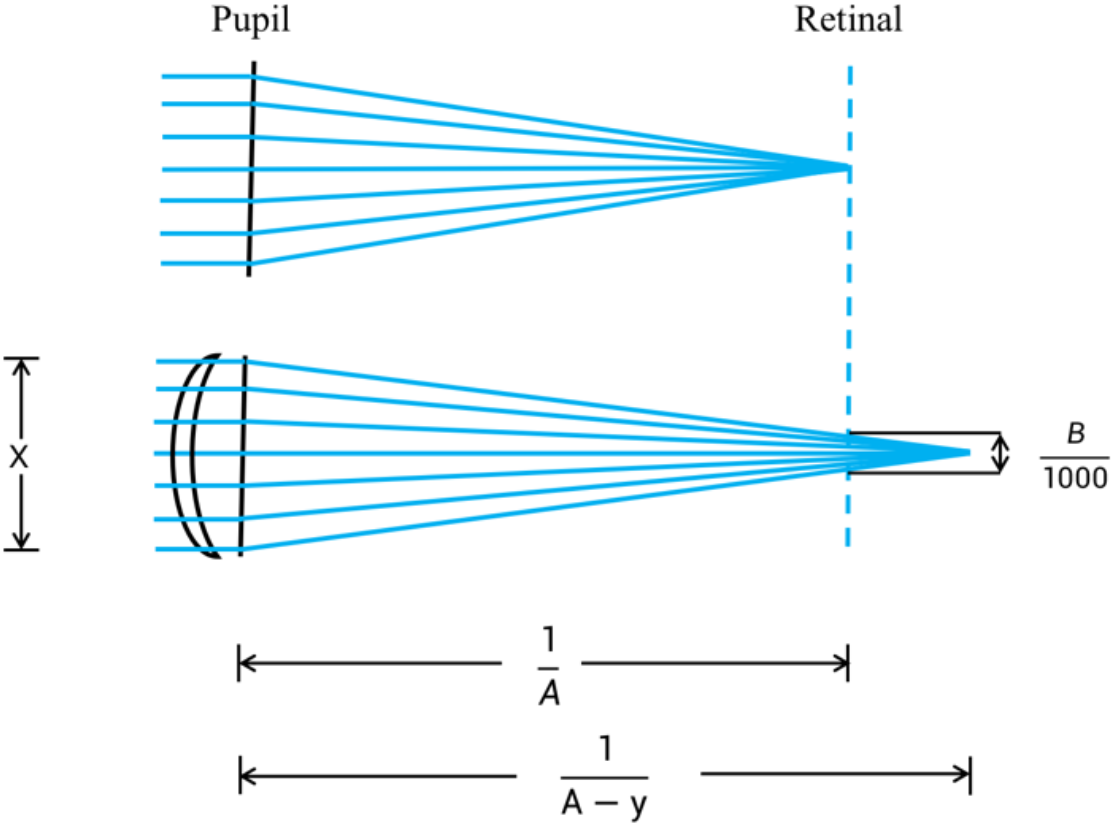
The optical model of minimum limit of discernible change in spherical power

By substituting the total diopter of human eye, the diameter of cone cell and the pupil diameter in Formula (4), we can derive the resolution limit of human eye to spherical lens change. Since the value of y is very small, the influence of the vertex distance on the Formula (1) can be ignored

### Clinical validation of the resolution limit of human eye to spherical lens change

#### Study participants

The data were prospectively collected from myopia volunteers who visited Beijing Tongren Hospital from September 2020 to September 2021. The research was approved by the Human Studies Committee at Beijing Tongren Hospital (Beijing, China) in accordance with the Code of Ethics of the World Medical Association (registration number: ChiCTR2100047074). Subjects signed a statement of informed consent prior to their participation in the study. Patient’s inclusion criteria were as follows: (1) patients with myopia not more than -6D, with corrected visual acuity≧ 1.0; (2) those having astigmatism with rule and ≤0.25D; and (3) those having good compliance and able to complete the optometry, and red-green balance check. Exclusion criteria were as follows: (1) Patients complicated with heterotropia, amblyopia or ocular infection and other eye diseases; (2) those with a history of ocular trauma; (3) those undergoing surgery or other corrective therapy; and/or (4) those with refractive errors caused by genetic or congenital factors.

### Examination procedures

#### Adjust the spherical power at an interval of 0.25D to find the red-green balance point

Standard subjective refraction was performed using a standard phoropter. The monocular best vision sphere was determined using the Duochrome chart to ensure the circle of least confusion was on the retina before conducting Jackson Cross Cylinder (JCC). The circle of least confusion was maintained on the retina as cylinder power was increased. Once cylinder power and axis were calculated, the sphere was refined to best-corrected visual acuity (BCVA)with minimum minus power. Binocular balance using alternate occlusion technique was then conducted to best binocular VA to BCVA with minimum minus power.

Binocular balanced refraction results were placed in a trial frame and the eye not being tested was occluded. Subjects were asked to give a response as to whether the letters on the red or green side of the Duochrome were clearer. If the subject reported the red side being clearer, minus power was added in 0.25D steps until equality between red and green was achieved. If the subject reported the green side being clearer, then plus 0.75D was added so that red became clearer. The same step as previously described for when the side was clearer was then performed. The final sphere was where equality was first achieved between the red and green letters of the Duochrome. If equality could not be achieved, then the final sphere was the point where the next 0.25D change made green clearer. The final sphere, and previously calculated cylindrical power and axis were recorded for the final prescription.

#### Verify the resolution limit of change in spherical power

Starting from the diopter of red-green balance point of equal clarity obtained by adjusting the spherical power at an interval of 0.25D, the diopter of spherical power was increased and decreased by one time of resolution limit, respectively. The volunteers were asked for any changes in the clarity of the red and green optotypes either from equal clarity to green clarity or red clarity. If there was no change, the point of equal clarity was used as the starting point again, and the diopter of spherical power was changed by 2 times the resolution limit. The volunteers were then asked for any changes in the clarity of optotypes. If still no change, the point of equal clarity was taken as the starting point to further increase the range of changes in the diopter of spherical power (an integer multiple of the resolution limit) until the clarity of optotypes changed.

In case of failure to identify the red-green balance point of equal clarity by adjusting the spherical power at an interval of 0.25D, the maximum diopter of red clarity (an increase of -0.25D to be green clarity) should be taken as the starting point, and the resolution limit as the interval to find the point of equal clarity. After identifying the point of equal clarity, the limit of differences in the visually resolved diopter of spherical power could be validated following the above steps. If the point of equal clarity was still not found at the interval of the resolution limit, the diopter of spherical power was increased by one time the resolution limit from the maximum diopter of red clarity to get green clarity, and the volunteer’s resolution limit was denoted as 1/2 of the resolution limit.

The prescription and the best corrected visual acuity were obtained by adjusting the spherical power at an interval of 0.25D and the resolution limit, respectively. The prescription of spherical power was determined according to the following principles: In case of a red-green balance point, the diopter of equal clarity is taken as the prescription diopter of spherical power. In absence of the red-green balance point, the maximum diopter of red clarity is taken as the prescription diopter of spherical power. The two sets of trial case lenses were of the same substrate material, manufacturing process and manufacturer (Minghao Technology (Beijing) Co., Ltd.). The visual acuity chart was an international standard one, and the recorded values were converted to the logarithm of the minimum angle of resolution (LogMAR).

### Statistical analysis

Before study initiation, we calculated the necessary sample size to ensure feasibility, we recruited 30 volunteers to observe the resolution limit of discernible change in spherical power. The mean resolution limit of change in spherical power was 0.05±0.01. The test level was 0.05 and the test power was 0.80, 110 volunteers were needed based on the following sample size calculation formula. (*δ*=allowable error,α=inspection level, *σ*=standard deviation)

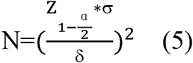

SPSS 20.0 (IBM, Armonk NY) statistical software was used for analysis. Statistical significance was set at p≤0.05. For continuous variables, data are presented as mean ± SD. Distributional normality was tested between different groups. The single-sample t-test was used to compare the theoretically derived resolution limit and the actual resolution, and the paired *t*-test was used to test the comparison of the spherical equivalent prescription and corrected visual acuity.

## Results

### The theoretically derived resolution limit of human eye to spherical lens change

When the total diopter of a human eye was 60.00D, the diameter of cone cell of 3 μm^[4]^, and pupil diameter of 4 mm, they were substituted into Formula (4). The theoretically derived resolution limit was 0.05D, and the manufacturing interval of new spherical-power trial case lenses in clinical validation was set at 0.05D.

### Clinical validation of the theoretically derived resolution limit of human eye to spherical lens change

As shown in Table 1, a total of 119 volunteers were enrolled in this study, with an average age of 23.7±2.3 years. The probability of achieving red-green balance when the spherical power was adjusted at an interval of 0.25D was much lower than that when the spherical power was adjusted at 0.05D.

**Table 1.** Basic information of included volunteers

|  | Right eye | Left eye |
| --- | --- | --- |
| Gender |  |  |
| Male | 48 |  |
| Female | 71 |  |
| Age | 23.7±2.3 |  |
| Red-green balance rate of the spherical power adjusted at an interval of 0.25D | 17.6% (21 eyes) | 12.6% (15 eyes) |
| Red-green balance rate of the spherical power adjusted at an interval of 0.05D | 84% (100 eyes) | 84% (100 eyes) |

The resolution limit that volunteers can actually resolve is shown in Table 2. Since the volunteers were myopic, increasing the diopter of spherical power from the point of equal clarity was equivalent to undercorrection, and the volunteers were required to observe whether the red optotypes became clearer. Whereas, decreasing the diopter of spherical power from the point of equal clarity was equivalent to overcorrection, and the volunteers were required to observe whether the green optotypes became clearer. It can be seen from the results that there is no difference between the theoretically derived value and the actual resolution when the diopter of spherical power is increased, but of difference when the diopter of spherical power is decreased. The right eye (t=-4.206, P=0.000) and left eye (t=-2.993, P=0.003) were more likely to detect the changes in the diopter of spherical power on a red background.

**Table 2.** Limit of discernible change in spherical power

|  | Actual resolution<br>(D) | Theoretically<br>derived value (D) | t | P |
| --- | --- | --- | --- | --- |
| Increase in diopter of spherical<br>power (undercorrection) |  |  |  |  |
| Right eye | 0.05±0.01 | 0.05 | -1.679 | 0.096 |
| Left eye | 0.05±0.02 | 0.05 | -0.911 | 0.364 |
| Decrease in diopter of<br>spherical power<br>(overcorrection) |  |  |  |  |
| Right eye | 0.06±0.04 | 0.05 | 3.336 | 0.001 |
| Left eye | 0.06±0.03 | 0.05 | 2.334 | 0.021 |

As shown in Table 3, 98.3% of the right eyes in volunteers could resolve the changes in 0.05D of spherical power with the addition of the spherical power and 78.9% with the decrease of the spherical power; and 96.7% of the left eyes could resolve the changes in 0.05D of spherical power with the addition of spherical power and 83.2% with the decrease of spherical power.

**Table 3.** Distribution of the resolution limits for changes in the actually resolved diopter of spherical power

|  | OD |  | OS |  |
| --- | --- | --- | --- | --- |
|  | Increase in<br>spherical lenses<br>(%) | Decrease in<br>spherical lenses<br>(%) | Increase in<br>spherical lenses<br>(%) | Decrease in<br>spherical lenses<br>(%) |
| 0.025D | 16 (13.4%) | 16 (13.4%) | 19 (16.0%) | 19 (16.0%) |
| 0.05D | 101 (84.9%) | 78 (65.5%) | 96 (80.7%) | 80 (67.2%) |
| 0.10D | 1 (0.8%) | 22 (18.5%) | 2 (1.7%) | 17(14.3%) |
| 0.15D | 1 (0.8%) | 2 (1.7%) | 2 (1.7%) | 2 (1.7%) |
| 0.20D | 0 (0.0%) | 1 (0.8%) | 0 (0.0%) | 0 (0.0%) |
| 0.25D | 0 (0.0%) | 0 (0.0%) | 0 (0.0%) | 1 (0.8%) |
| Total | 119 (100.0%) | 119 (100.0%) | 119 (100.0%) | 119 (100.0%) |

Table 4 shows the prescription spherical equivalent and corrected visual acuity when the spherical power are adjusted at intervals of 0.25D and 0.05D. The 0.25D group was undercorrected, and the spherical equivalent and corrected visual acuity were significantly lower than those in the 0.05D group.

**Table 4.**
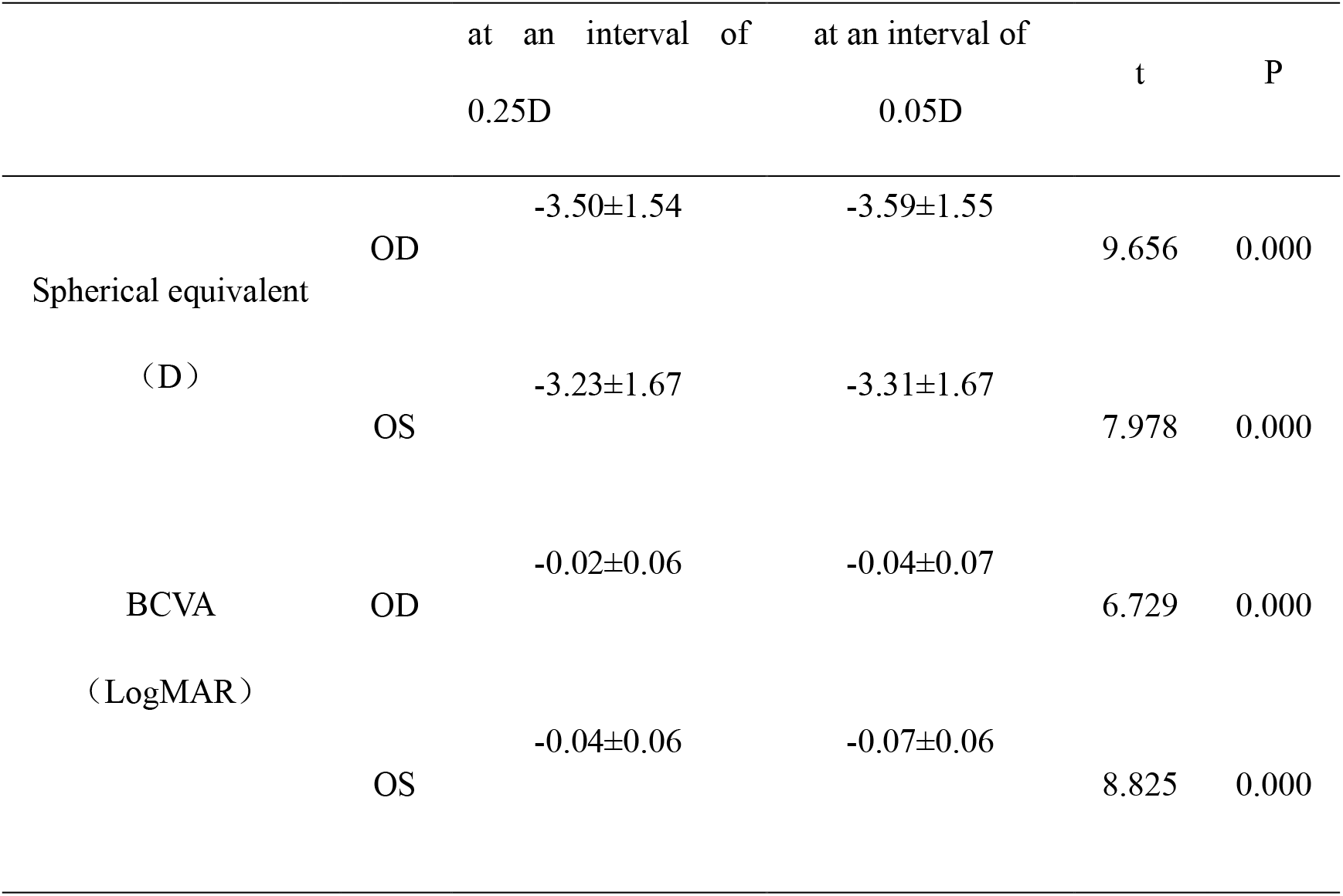
Prescription and corrected visual acuity for adjusting spherical power at 0.25D and 0.05D

## Discussion

In 1865, the diopter was defined as the unit of optometry (at 1D interval) at an international ophthalmology conference in Heidelberg^[7]^. In 1900, the trial lenses at an interval of 0.25D appeared, greatly improving the effect of rectifying ametropia with eyeglasses. With the popularization of resin lens and computer numerical control (CNC) free-form surface machining, the machining accuracy of lens has been greatly improved from that 100 years ago and we can produce lenses at intervals of less than 0.25D nowadays.

Optometry aims to determine the lens power needed to achieve the BCVA. This power is defined by forming a circle of least confusion (COLC) in the macula, so it depends critically on the focusing accuracy of optical lens and the subject’s ability to identify test-objects of different clarities. The diopter value of ametropia is a continuous variable and the subjective optometry is to simulate the continuous variable by inserting trial lenses with different powers into a trial frame. When the diopter value is reached, the external parallel rays of light entering the eye form a COLC, thereby obtaining the BCVA. COLC cannot be formed in case of any difference between the variable and the power of inserted lenses, so the BCVA also depends on the resolution limit of discernible change in lens power. If the difference is less than the resolution limit, the BCVA will be obtained properly; otherwise, inaccurate value may be obtained. Therefore, adjusting the combination of trial lenses per resolution limit is relatively conducive to obtaining the BCVA.

The normal visual acuity changes with diameters of cone cells and pupils^[8 9]^. With the same optical model, we derive the formula for calculating the resolution limit of discernible change in spherical power. The diameters of cone cells in human eyes range from 0.5 um to 4 um^[10]^, and the pupil diameter ranges from 2 mm to 4 mm^[11]^in daytime, so the resolution limit theoretically ranges from 0.01D to 0.12 D. In this study, the cone cell diameter was assumed to be 3 um and the pupil diameter was 4 mm, thereby the resolution limit obtained was 0.05 D. If the cell diameter remains unchanged (3 um) and the pupil diameter changes from 3 mm to 4.5 mm, the resolution limit fluctuates between 0.06 D and 0.04 D.

When the COLC is formed by focusing on a test-object on monochrome background, it is often difficult to identify the change in test-object clarity due to lack of reference object. In 1927, Brown devised the red-green Duochrome test to solve the problem, which has been an important step of the SOP for optometry since 1950s^[12-14]^. For subjective optometry, the ideal result is that the visible light centered at a wavelength of about 570 nm is exactly focused on the retina, and the black characters on the red and green backgrounds should have the same clarity when the broad-band visible light passing through the corrective lens is focused on the retina^[15 16]^. In this study, to obtain more reliable results, the red-green balance test was used to assist volunteers in identifying changed clarity of the test-object after adjusting the spherical power.

As shown in Table 2, the theoretical resolution limit is in good agreement with the actually discernible diopter on the red background. Although the difference is only 0.01D, a significant difference is found between the theoretical resolution limit and the actually discernible diopter on the green background, which is related to the subjective preference of patients, which affects the results of the red-green balance test^[17 18]^. Study suggests that red test-object is easier to be perceived than green one^[19 20]^, so the background color also affects the activity of visual cortex. As shown in Table 3, more than 95% of people can discern a 0.05D change in spherical power on the red background, and about 80% of people can discern a 0.05D change in spherical power on the green background, which are consistent with the theoretical results. The discernible resolution limits of 13-16% volunteers are less than 0.05D, which may be attributed to their smaller cone cell diameter or larger pupil diameter.

As shown in Table 4, the spherical equivalent and the BCVA of left and right eyes in the 0.25 D interval group are significantly lower than those in the 0.05 D interval group and under correction. The situation can be attributed to the fact that the 0.25 D interval group could only set the minimum diopter achieving clear red test-object as the prescribed diopter when the red-green balance cannot be achieved.

The factors affecting the results of red-green balance test include eye accommodation, patients’ subjective preferences, cataract, pupil size and ambient brightness^[21-23]^. Our study shown that only 17.6% of right eyes and 12.6% of left eyes can achieve the red-green balance when spherical power is adjusted at an interval of 0.25D. These volunteers with cataract, abnormal pupil and other abnormalities had been excluded before test. After adjustment of the interval to 0.05D, the red-green balance rate of both eyes increasing to 84% in the same experimental environment indicates that the adjustment interval of spherical power is critical to the red-green balance. As shown in Table 2, the change in spherical power to turn clear test-object on red background to that on green background is about 0.11D. Accordingly, it is difficult to achieve the red-green balance by adjusting the spherical power at an interval of 0.25D, unless the power achieving red-green balance is an integral multiple of 0.25D. Liu et al.^[24]^ carried out a study on necessity of red-green balance test in subjective optometry and found there was no difference in results between the patients subject to red-green balance test and those free from the test. However, the spherical power was adjusted at an interval of 0.25D and different results may be obtained if the red-green balance rate is improved by adjusting the spherical power at an interval of 0.05D.

The major limitations of this study are that we only observed the short-term influence of different intervals on corrected visual acuity during optometry, so we will continue to explore the long-term influence of eyeglasses made with optometry prescription obtained by adjusting spherical power at an interval of 0.05D on binocular balance, wearing comfort and myopia progression as there is evidence that full correction are beneficial for myopia control than under correction^[2 25 26]^.

In summary, this study is the first one that deduce theoretically and verify the resolution limit of human eye to spherical lens change. The results show that different intervals of trial lens significantly influence the results of the refraction and Duochrome test. The 0.05 D interval group produces a more accurate refraction and a better visual acuity, which is linked to a higher rate of red-green balance. This study provides a great potential for future improvement of refraction technique and accuracy.

## Data Availability

All data produced in the present study are available upon reasonable request to the authors

## ACKNOWLEDGEMENTS

The authors acknowledge the support from Beijing Advanced Innovation Center for Big Data-Based Precision Medicine. The authors also thank Wenhua Wang, Hengheng Tang, Weiwei Hao for their help of investigation.

## Source of Funding

The authors acknowledge the support from the Beijing Advanced Innovation Center for Big Data-Based Precision Medicine and Beijing Municipal Science and Technology Project (Z201100005520042)

## Conflict of Interest

The authors declare no conflict of interest.

## Notes

### Competing Interest Statement

The authors have declared no competing interest.

### Clinical Trial

ChiCTR2100047074

### Author Declarations

The research was approved by the Human Studies Committee at Beijing Tongren Hospital (Beijing, China) in accordance with the Code of Ethics of the World Medical Association (registration number: ChiCTR2100047074).

